# Cohort profile: the Cohort for Risk Prediction Model Evaluation (CORE) for external validation of models identifying high-risk pregnant women in the early second trimester

**DOI:** 10.64898/2026.07.02.26357113

**Authors:** Rahul Jain, Nikhil Sharma, Ashok Khurana, Nitya Wadhwa, Reva Tripathi, Abhinav Jain, Shinjini Bhatnagar, Ramachandran Thiruvengadam, Bapu Koundinya Desiraju

**Author notes:** **Corresponding author:** Dr. Bapu Koundinya Desiraju, Translational Health Science and Technology Institute, National Capital Region Biotech Cluster, 3rd Milestone, Gurgaon-Faridabad Expressway, Faridabad, India, 121001).

## Abstract

**Purpose:** The Cohort for Risk Prediction Model Evaluation (CORE) was established to externally validate prediction models that identify high-risk pregnancies in the early second trimester. Such models are often developed on small, single-source datasets and seldom tested elsewhere. Recent evidence shows that only about 6–10% models are ever externally validated which raises concerns about whether they perform reliably in new and diverse populations. In the maternal–perinatal space, CORE addresses this gap by providing an Indian second-trimester cohort with harmonised imaging and outcome data on which existing risk prediction models can be validated.

**Participants:** CORE includes 964 pregnant women aged over 18 years, enrolled at the Hamdard Institute of Medical Sciences and Research (HIMSR), New Delhi, between August 2021 and March 2023. Women were recruited before 20 weeks of gestation and followed up at 18–22 weeks for an ultrasound scan and at delivery. At all time points, a structured set of sociodemographic, clinical, and obstetric data was captured, together with ultrasound images at 18–20 weeks from which fetal biometry and cervical length were measured.

**Findings to date:** The median maternal age was 27.6 years; 51% had a normal body-mass index (BMI) and 30% were overweight. There were almost equal number of Nulliparous (480, 50%) and multiparous (484, 50%).About 41% prevalence of history of prior preterm in multiparous women. Outcomes were available for 750 participants (23 abortions, 3 stillbirths, 724 singleton live births). Among the 724 live births, 80/724 (11%) were preterm, 190/716(26.5%) were small for gestational age (SGA) and 40/716 (5.6%) were large for gestational age (LGA) by INTERGROWTH-21st standards and 260/716(35.6%) of newborns were categorized into small vulnerable newborn(SVN).

**Future plans:** CORE will be used to externally validate and, in aggregate with similar cohorts, help improve risk-prediction models for pregnant women in India and comparable settings. We invite collaborators to use this resource; clinical and imaging data are available under a controlled-access model on reasonable request.

**Strengths and limitations of this study:**

- Prospective cohort with data-collection and ultrasound protocols harmonised with the GARBH-Ini and AMANHI cohorts, enabling like-for-like pooled and cross-cohort analyses.
- A two-tier ultrasound quality-assurance process, with retention of both clean (unannotated) and caliper-annotated images, supports validation of image-based prediction models.
- Sample size informed by precision-based guidance for the external validation of prediction models, providing adequate power to assess model discrimination for the principal outcomes.
- Single-site, hospital-based recruitment from a limited geographical catchment, which constrains direct generalisability and makes the cohort most valuable when pooled with comparable cohorts.
- No continuous follow-up between 20 weeks of gestation and delivery, limiting the assessment of temporal change and longer-term outcomes.

## Introduction

Neonatal mortality and morbidity remain pressing global health concerns. Preterm birth, small for gestational age (SGA), and low birthweight (LBW) together account for a large share (1.4 million, approx 51 %) of neonatal deaths and stillbirths [1]. Beyond the immediate risks to the newborn, these conditions contribute to developmental difficulties and chronic ill-health across the life course, representing a substantial loss of human capital [2]. Despite decades of progress in health care, reductions in neonatal mortality have stagnated, particularly in low- and middle-income countries (LMICs). India, among the largest contributors to the global burden, faces challenges in meeting Sustainable Development Goal (SDG) 3.2, which targets the reduction of neonatal and under-5 mortality [3]. This underscores the need for strategies that identify and manage high-risk pregnancies early enough for timely intervention.

Despite repeated efforts to develop clinical prediction models for high-risk pregnancy, evaluation of these models beyond their development cohorts remains sparse. This is part of a wider pattern in clinical artificial intelligence (AI): machine-learning models are prone to overfitting on their training data, yet external validation is the exception rather than the rule. A systematic review of 516 studies reporting AI for diagnostic medical-image analysis found that only about 6% had undergone external validation [4]; a scoping review of AI pathology models for lung-cancer diagnosis reached a similar conclusion, with external validation reported in only a small minority of studies [5]. In critical care, where validation is comparatively mature, a recent meta-analysis showed that external validation is concentrated on a handful of public datasets such as MIMIC [6] and eICU [7], with model performance typically degrading on truly external data [8]. The same fragility is documented for prediction models in general, where external validation is infrequent and discrimination is usually worse than reported [9], and specifically in obstetrics, where maternal-characteristic models for spontaneous preterm birth have not reproduced their reported performance on independent external validation [10].

The limiting step now is not another model, but rigorously validated models that are safe to take toward clinical deployment. Progress depends on the availability of independent, well-characterised cohorts on which existing models can be tested. Individually, such cohorts are often modest in size and drawn from single sites; pooled across settings, however, they constitute a robust, distributed external-validation platform. CORE is designed to be one node in that platform. To this end, we describe the development of CORE, a cohort of pregnant women established at a tertiary-care hospital in India and designed specifically for the external validation of models that identify high-risk pregnancies in the early second trimester. By assembling harmonised clinical, imaging, and demographic data, CORE provides a real-world LMIC platform for assessing model performance. We do not present CORE as a complete solution to the external-validation gap. Rather, it is a step in the right direction: an LMIC-based resource on which models can be tested and, in aggregate with similar cohorts, refined for heterogeneous populations.

## Cohort description

### Study design

This is a prospective observational cohort study of pregnant women (N = 964) conducted at the Hamdard Institute of Medical Sciences and Research (HIMSR), a tertiary-care hospital in New Delhi, India, from August 2021 to March 2023. Women were enrolled at < 20 weeks of gestation and followed up at the 18–20 week scan and at delivery.

This manuscript was prepared in accordance with the Strengthening the Reporting of Observational Studies in Epidemiology (STROBE) guidelines for cohort studies, and the completed STROBE checklist is provided as a supplementary file.

### Ethical approval and informed consent

The study was approved by the institutional ethics committees of HIMSR (HIMSR/IEC/43/2021) and the Translational Health Science and Technology Institute (THSTI), Faridabad (THS 1.8.1/(89). Written informed consent was obtained after the nature of the study was explained. For illiterate participants, a thumb impression was taken after the participant confirmed understanding and gave explicit verbal consent, and a literate impartial witness countersigned the consent form.

### Participants

Inclusion criteria were age > 18 years, a single live intrauterine pregnancy, gestational age estimated from crown–rump length (CRL) measured before 14 weeks of gestation, and < 20 weeks of gestation at enrolment. Gestational age for enrolment, scheduling of follow-up visits, and outcome evaluation was calculated from the CRL measured at < 14 weeks using the Hadlock formula[11]. A total of 964 pregnant women who attended HIMSR for antenatal care, met the inclusion criteria, and provided consent were enrolled.

### Clinical and sociodemographic data at enrolment

Information on social factors - type of household, educational status of the participant and the head of the family, occupation, family income, and religious faith - was collected. Demographic details such as the type of cooking fuel, source of drinking water, and family size were recorded, along with lifestyle factors including habitual smoking and tobacco chewing. Clinical data such as age, height, and weight were obtained from hospital records. Past obstetric history -parity, number of live births, and any history of adverse outcomes such as preterm birth and abortion and their number was recorded. Data on the current pregnancy, including date of last menstrual period (LMP), interpregnancy interval, use of assisted conception, and menstrual irregularities, were documented. The full list of variables captured in each category is given in Table 1.

**Table 1:**
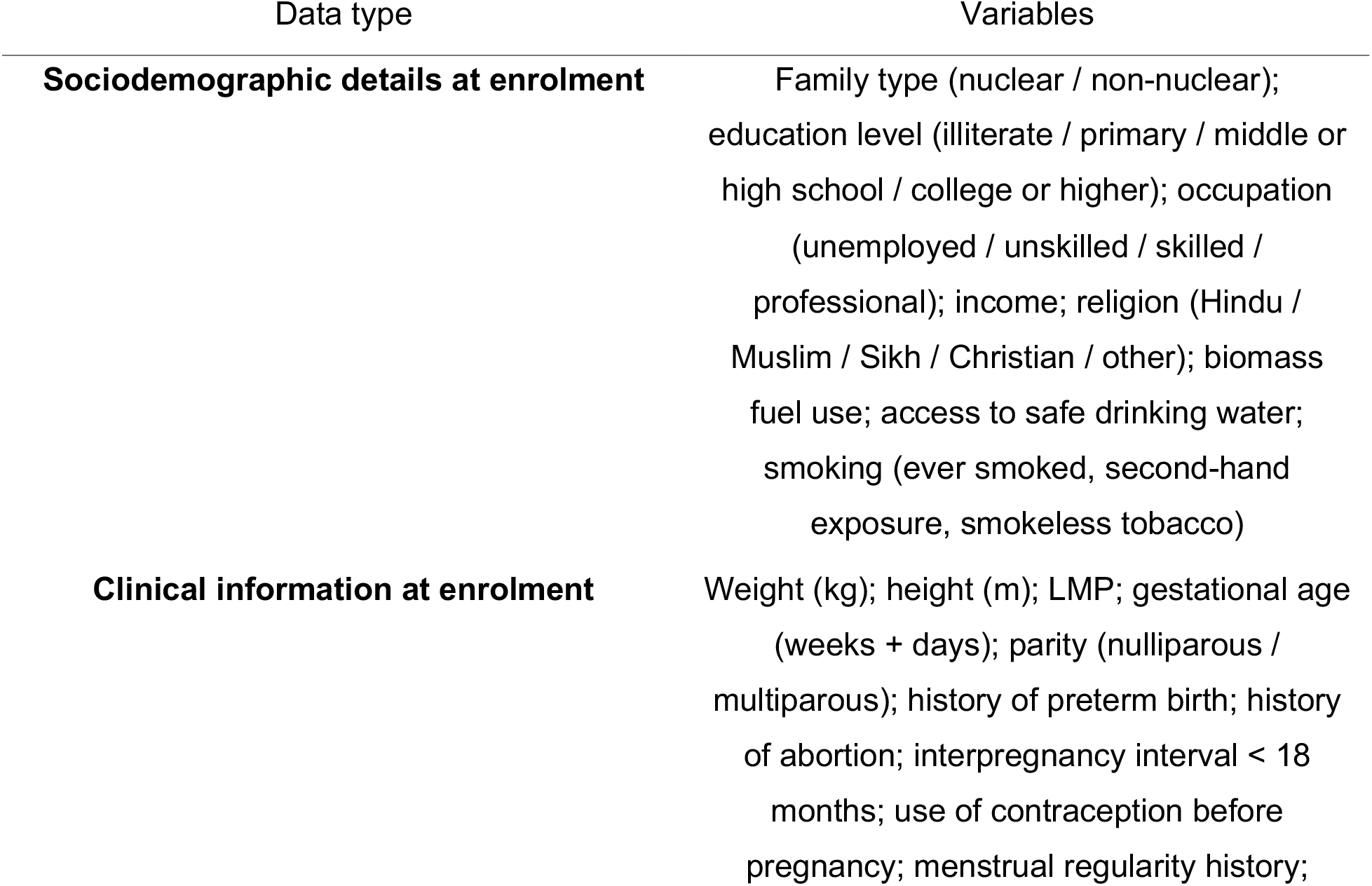
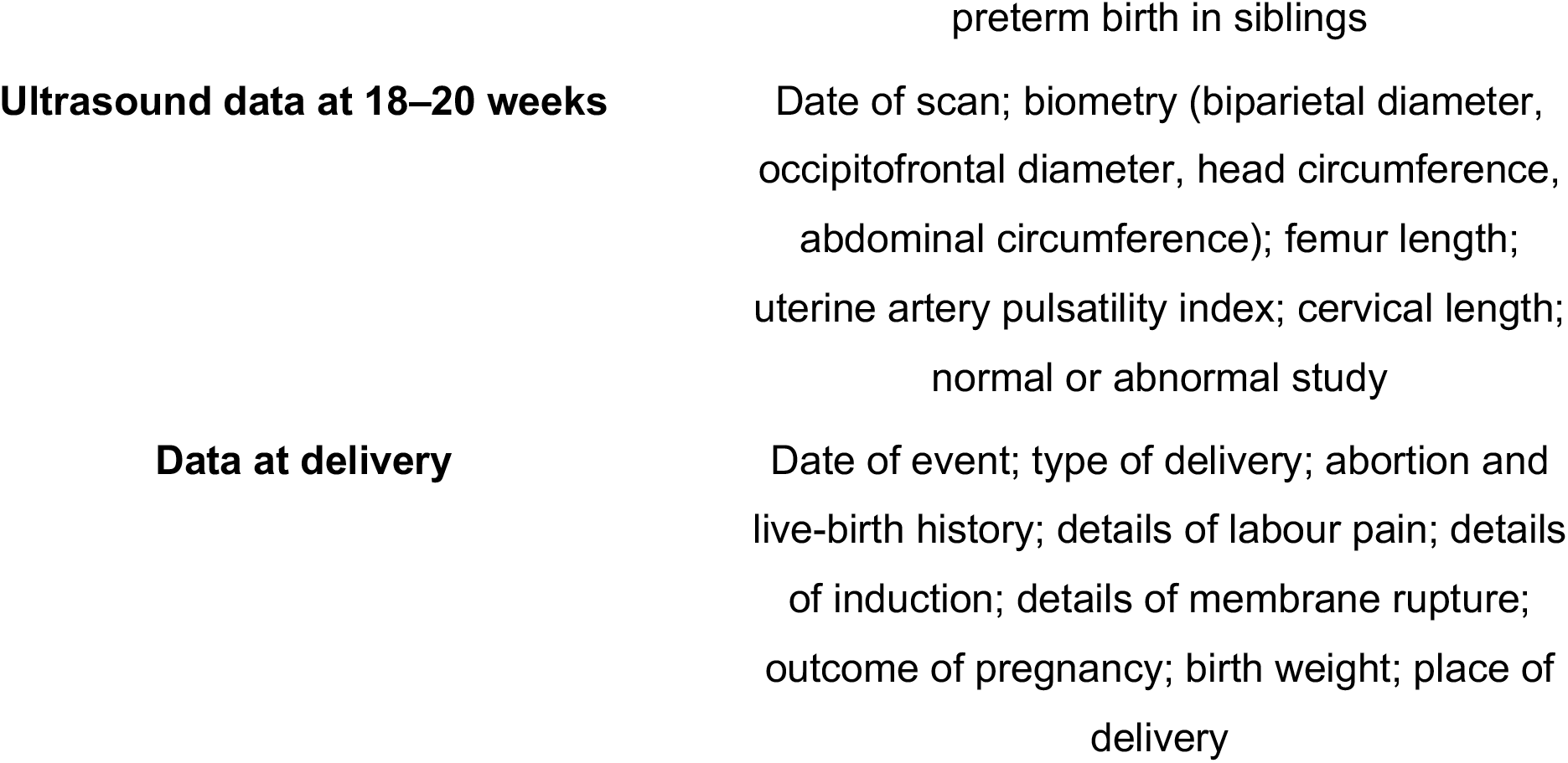
List of participant characteristics captured in the cohort study.

### Ultrasound data

A level-2 (anomaly scan) ultrasound scan was performed by a trained radiologist at 18–20 weeks of gestation on a Samsung HS70A .Images of the fetal head, femur, and abdomen were acquired in triplicate using a transabdominal approach, and three images of the maternal cervix were acquired transvaginally. Biometric parameters (head circumference, biparietal diameter, occipitofrontal diameter, abdominal circumference, femur length) and cervical length were measured from the acquired images. Cervical length was measured transvaginally [empty bladder, sagittal view, shortest of three measurements]. Colour Doppler was used to measure the uterine artery pulsatility index [protocol: bilateral measurement of left and right uterine arteries at the level of the internal cervical os, reporting mean PI, and documenting presence or absence of early diastolic notching on each side]. For each biometric parameter and the cervix, three images were captured without calipers (clean, unannotated images) and the same images were re-saved after caliper placement with annotations.

### Data management

Study data were collected and managed using the Research Electronic Data Capture (REDCap) tool hosted on a secure server at THSTI. REDCap is a secure, web-based platform designed to support data capture for research and is compliant with 21 CFR Part 11, FISMA, HIPAA, and GDPR. All ultrasound images were de-identified prior to transfer by cropping and removing personal identifiers, then classified into two categories (a) clean images without measurements and (b) labelled images containing measurements and annotations and stored separately. The two image sets were automatically transferred over a secure VPN connection to the image database at THSTI. All ultrasound protocols were harmonised with GARBH-Ini and AMANHI. The complete workflow for ultrasound image transfer is shown in Figure 2.

### Patient and public involvement

Patients and members of the public were not involved in the design, conduct, reporting, or dissemination plans of this research.

### Quality control and quality assurance

Rigorous quality control processes were in place for both clinical and ultrasound data collection. For clinical data, edit checks were built into REDCap, and a monitoring team at THSTI performed manual checks, identified discrepancies, and raised queries that were resolved by the clinical-site team using source documents. Ultrasound image quality assurance was a two-tiered process. In tier 1, the scanning radiologist evaluated three images per parameter and marked the best of the three. In tier 2, expert radiologists reviewed 10% of images monthly and scored them on a structured rubric — a 6-point scale for cervical images and a 7-point scale for biometry images, covering plane of image, zoom, and caliper placement as documented before.[12]

It was developed with stringent quality control and standardised protocols harmonised with other large pregnancy cohorts - the Interdisciplinary Group for Advanced Research on Birth Outcomes–DBT India Initiative (GARBH-Ini) [12] and the Alliance for Maternal and Newborn Health Improvement (AMANHI) [13], so that model inputs and outcomes can be compared like-for-like across cohorts.

### Outcomes

The events leading to delivery such as time of onset, nature of labour (spontaneous or induced) and rupture of membranes (prelabour or preterm premature) were recorded, together with the nature of delivery (vaginal, caesarean, assisted), reasons for augmentation and caregiver-initiated deliveries, and abortions, stillbirths, or intrauterine deaths. Birth weight, date of birth, and sex of the newborn were also documented. The full list of variables collected in each category is given in Table 1.

### Outcome definitions

**Preterm birth** was defined as a live birth at or after 22 0/7 weeks and before 37 0/7 weeks of gestation, consistent with the WHO definition [14]; pregnancy losses before 22 weeks were classified as abortions and reported separately. **SGA** (small for gestational age) was defined as a birth weight below the 10th percentile for gestational age and sex per the INTERGROWTH-21st standards [15]. **LGA** (large for gestational age) was defined as a birth weight above the 90th percentile for gestational age and sex per the same standards.

**SVN** (Small Vulnerable Newborn) was defined as per the Small Vulnerable Newborn Consortium[1] as a liveborn neonate who meets one or more of the following criteria: preterm birth (<37 completed weeks of gestation), small for gestational age (birthweight below the 10th percentile for sex and gestational age using the INTERGROWTH-21st newborn-size standards), low birthweight (<2,500 g), or any combination of these.

### Plan of statistical analysis

#### Sample size

At the planning stage, the approximate sample size was estimated using the rule of thumb that at least 100 outcome events are desirable for evaluating prediction models [16]. Based on an expected preterm-birth (PTB) prevalence of approximately 13%, the initial target was 800 participants. As the study progressed, loss to follow-up was higher than anticipated; to maintain an adequate number of outcome events, an additional 164 participants were enrolled, giving a total of 964.

In line with current guidance for the external validation of clinical prediction models, we additionally examined the precision of validation-performance estimates expected at the observed event count. For external validation, Riley and colleagues recommend targeting acceptable precision around the C-statistic, the calibration slope, the calibration intercept, [17]. To estimate an AUROC of 0.75 with a 0.10 targets precision for the c-statistics,about 400 participants for SVN, 472 for SGA, and 927 for PTB are required, and to estimate 0.20 targets precision for the calibration slope, at least 2796 participants for SVN, 3050 for SGA, and 4515 for PTB are required. Overall, our cohort is sufficiently powered to evaluate discrimination of existing prediction models around an AUROC of 0.75 for all three outcomes, while calibration can be evaluated with more limited precision.

### Measures of estimates

Continuous variables are presented as median and interquartile range (IQR); categorical variables as counts and percentages (n, %), with denominators reported alongside each estimate. Missing data were not imputed; analyses for each variable used complete cases, and the number of participants with available data is given in the footnotes to Tables 2 and 3.

**Table 2:**
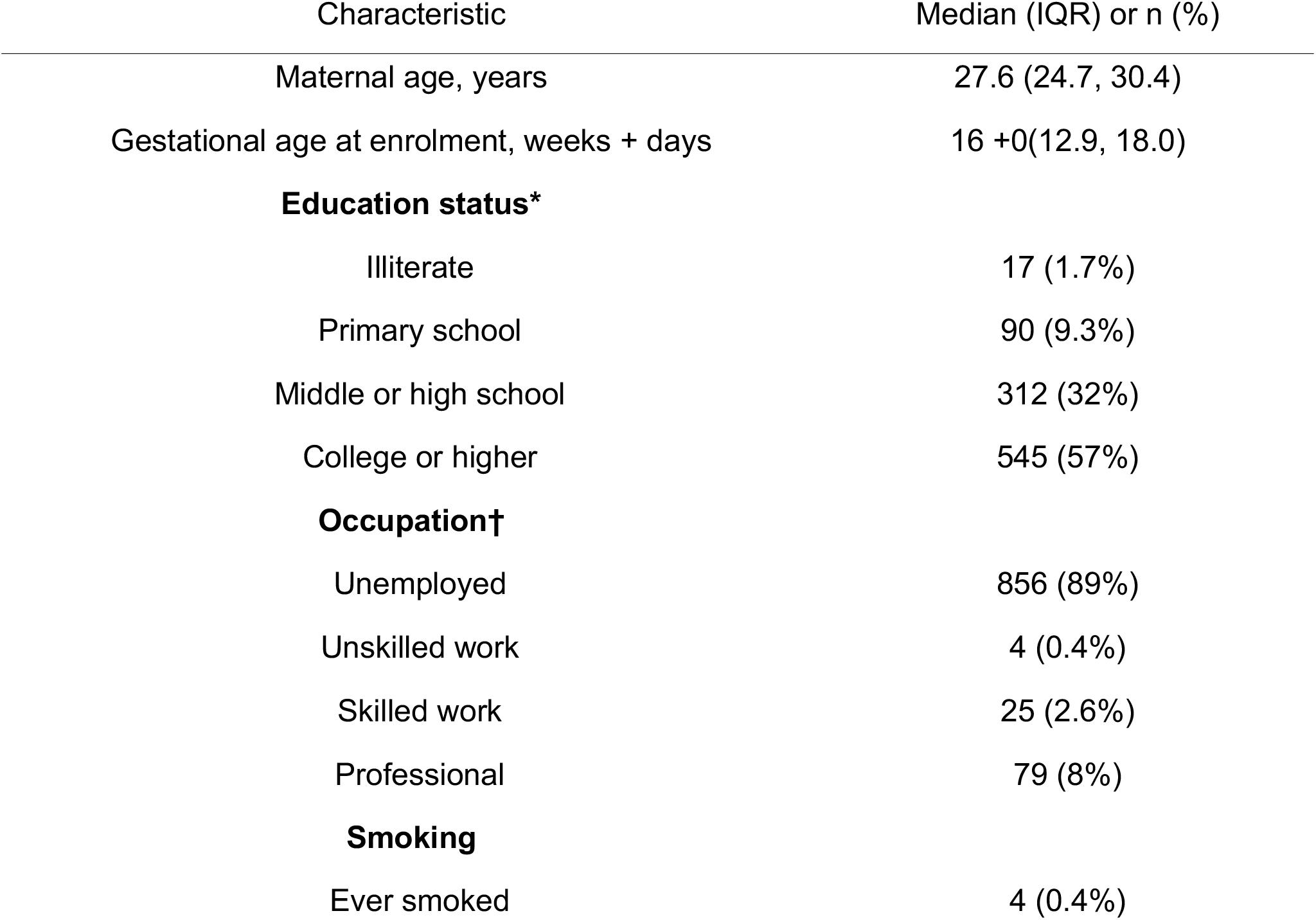
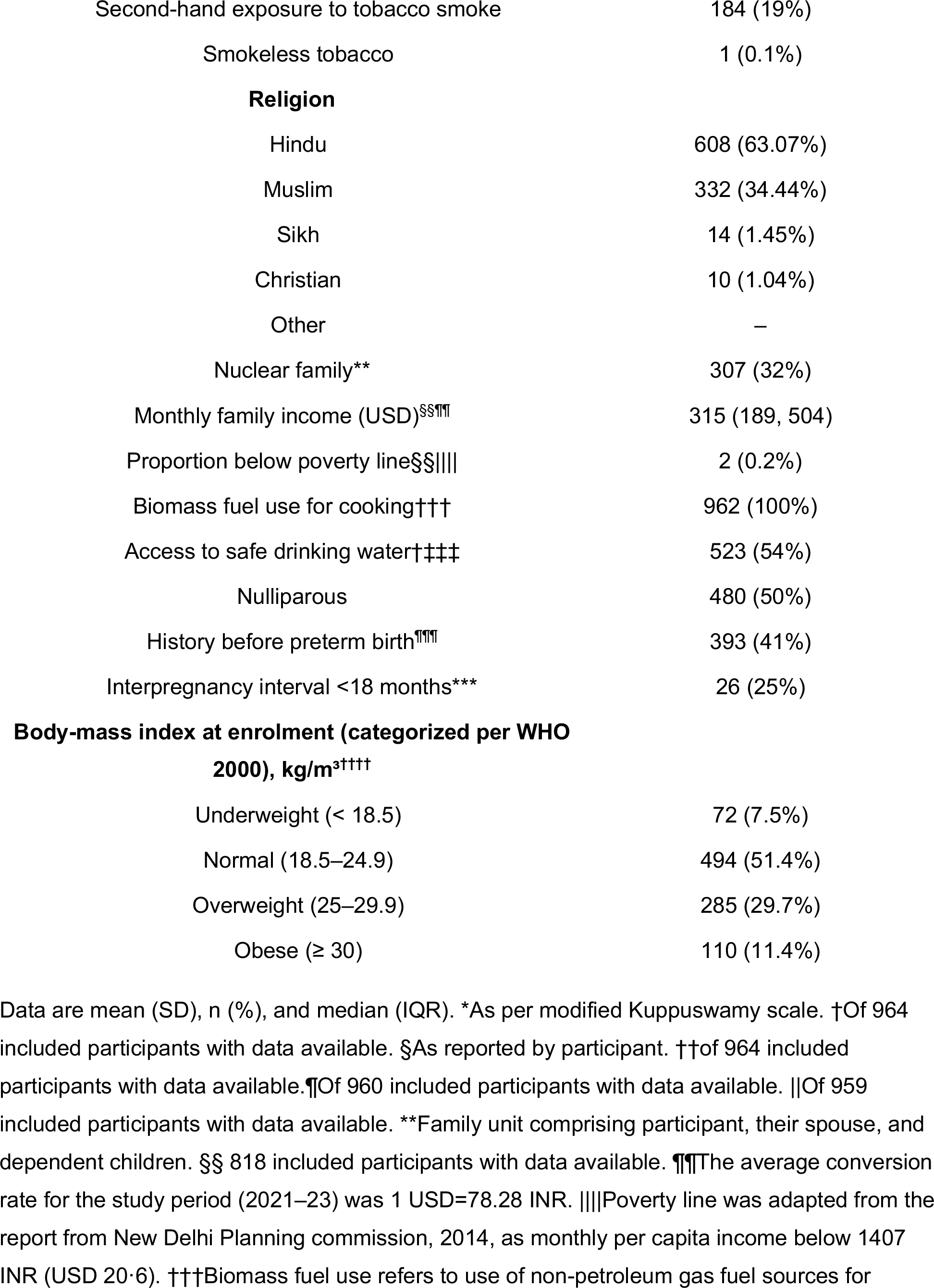
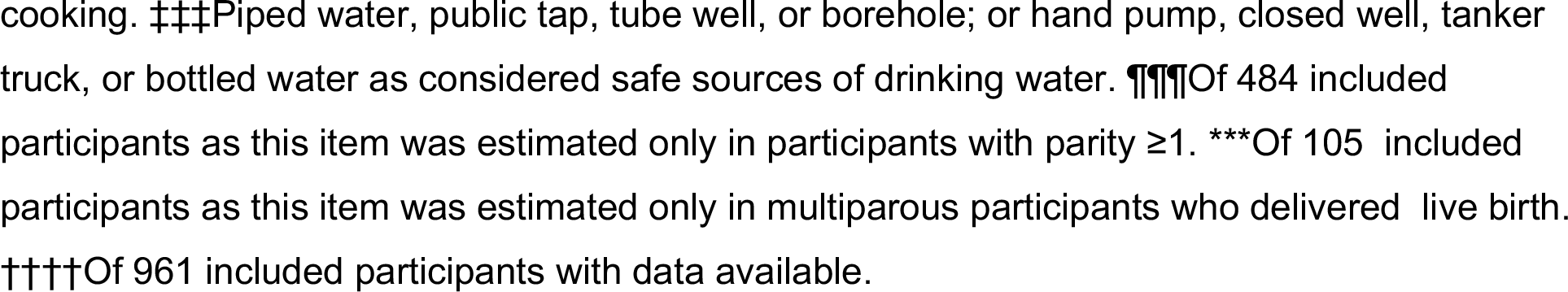
Sociodemographic and clinical characteristics of participants in the CORE cohort.

**Table 3:** Prevalence of outcomes in the cohort.

| Outcome | n/N | Prevalence % (95% CI) |
| --- | --- | --- |
| Preterm<sup>a</sup> | 80/724 | 11 (8.8–13.3) |
| SGA<sup>b</sup> | 190/716 | 26.5 (23.3–29.8) |
| LGA<sup>c</sup> | 40/716 | 5.6 (4.0–7.6) |
| Preterm–SGA | 12/716 | 1.7 (0.9–2.6) |
| Preterm–nonSGA | 67/716 | 9.4 (7.3–11.7) |
| Term–SGA | 178/716 | 24.9(21.8–28.3) |
| Term–nonSGA | 459/716 | 64 (60.7–67.5) |
| SVN<sup>d</sup> | 260/716 | 36.3(32.5–40.2) |

<sup>a</sup>Preterm birth: a delivery at or after 22 0/7 weeks and before 37 0/7 weeks of gestation. <sup>b</sup>SGA: birth weight below the 10th percentile for gestational age and sex per INTERGROWTH-21st standards [14]. <sup>c</sup>LGA: birth weight above the 90th percentile for gestational age and sex per the same standards. <sup>bc</sup> Out of 716 newborns with both birthweight and gestational age information available. <sup>d</sup>**SVN** (Small Vulnerable Newborn) was defined as per the Small Vulnerable Newborn Consortium as a liveborn neonate who meets one or more of the following criteria: preterm birth (<37 completed weeks of gestation), small for gestational age (birthweight below the 10th percentile for sex and gestational age using the INTERGROWTH-21st newborn-size standards), low birthweight (<2,500 g), or any combination of these.

### Studies and findings to date

Of 964 enrolled women, 754 (78.2%) underwent an ultrasound scan at 18–20 weeks, and outcomes were documented for 750 (77.8%) - comprising 23 abortions, 3 stillbirths, and 724 singleton live births. The flow of participants through the study is shown in Figure 1 and the baseline characteristics are summarized in Table 2. Participants had a median age of 27.6 years (IQR 24.7, 30.4). Approximately half (494, 51.4%) had a normal BMI, while a substantial proportion were overweight (285, 29.7%) or obese (110, 11.4%) and a small proportion were underweight (72, 7.5%).

**Figure 1:**
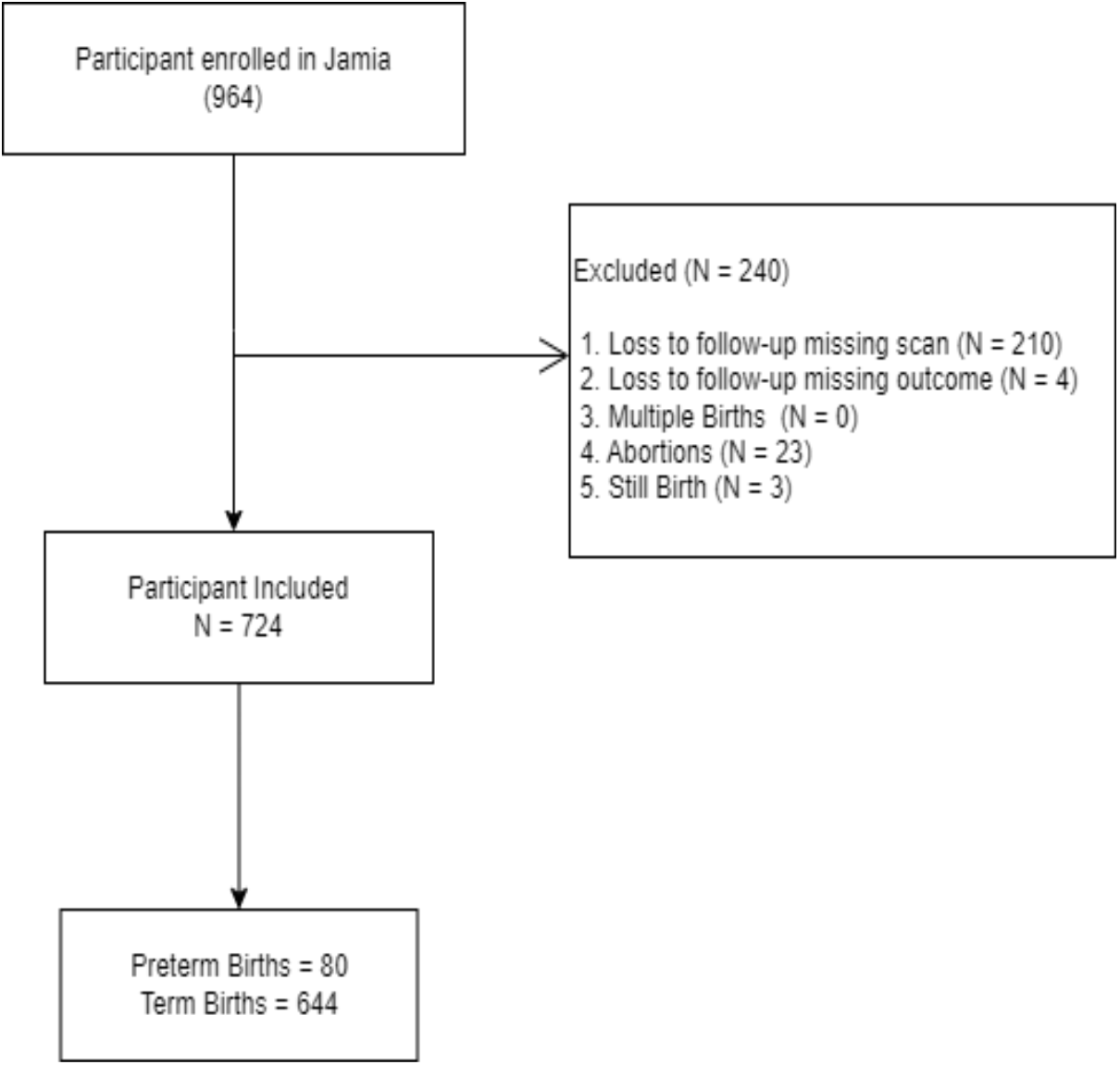
Study flow diagram summarizing participant enrolment, exclusions, and births included in the analysis.

**Figure 2:**
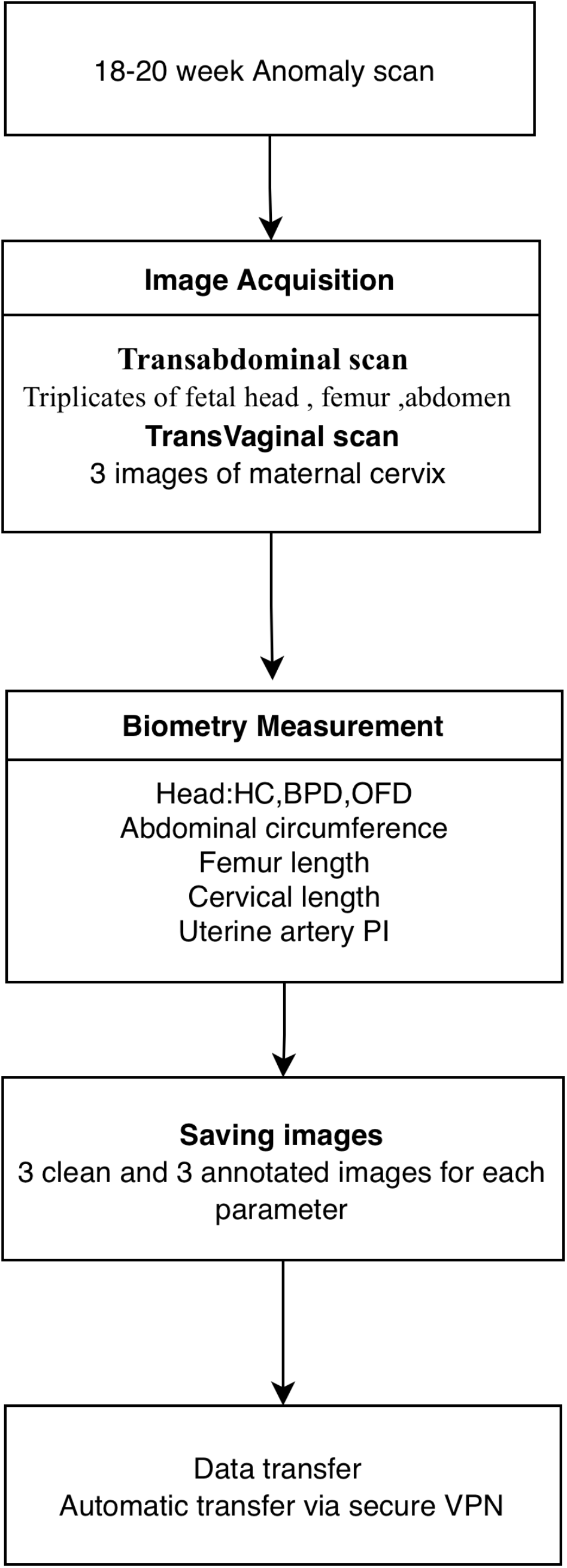
Workflow for de-identification, classification, and secure transfer of ultrasound images from HIMSR to the THSTI image database. Clean unannotated images and caliper-annotated images are stored separately and transferred over a secure VPN connection.

Most women were unemployed (856, 89%) and lived in non-nuclear families (657, 67%), with a median monthly family income of Rs. 25,000 (IQR 15,000, 40,000). Few reported smoking (4, 0.4%) or chewing tobacco (1, 0.1%), while a considerable number (184, 19%) were exposed to passive smoking. Almost all families (962, 99.8%) used LPG or biogas for cooking, and roughly equal proportions used safe (523, 54%) or unsafe (441, 46%) water sources.

The median gestational age at enrolment was 16 +0 (12.9, 18.0). Nulliparous (480, 50%) and multiparous (484, 50%) women were present in almost equal numbers. Among multiparous women, the most common prior risk factors were a history of past preterm births (41%), followed by short interpregnancy interval (31%), assisted conception (3.8%), and short cervical length (1.5%).

Among the 750 participants with recorded outcomes (23 abortions, 3 stillbirths, and 724 singleton live births), preterm birth occurred in 80/724 (11.0%), while 23/750 (3.1%) pregnancies ended in abortion and 3/750 (0.4%) in stillbirth.Of the 80 preterm births, 58 (72.50%) were late preterm (34–36 weeks), 17 (21.25%) moderately preterm (32 to < 34 weeks), 4 (5%) very preterm (28 to < 32 weeks), and 1 (1.25%) extremely preterm (22 0/7 to < 28 weeks). By phenotype, 30 (37.50%) followed preterm premature rupture of membranes, 19 (23.75%) followed spontaneous preterm labor, and 31 (38.75%) were caregiver-initiated. There were 101/716(14.1%) newborns with low birth weight (< 2.5 kg); the remaining 615/716 (85.8%) had a normal birth weight. Using the Small Vulnerable Newborn (SVN) definition, 260 of 716 newborns (36.3%) were classified as SVN. In the distribution presented in Table 3, 12/716 newborns (1.7%) were preterm and SGA, 67/716 (9.4%) were preterm and non-SGA, and 459 (64.1%) were term and non-SGA. The prevalence of the principal study outcomes, with 95% confidence intervals, is given in Table 3; the distributions of gestational age at delivery and of birth weight are shown in Figure 3.

**Figure 3:**
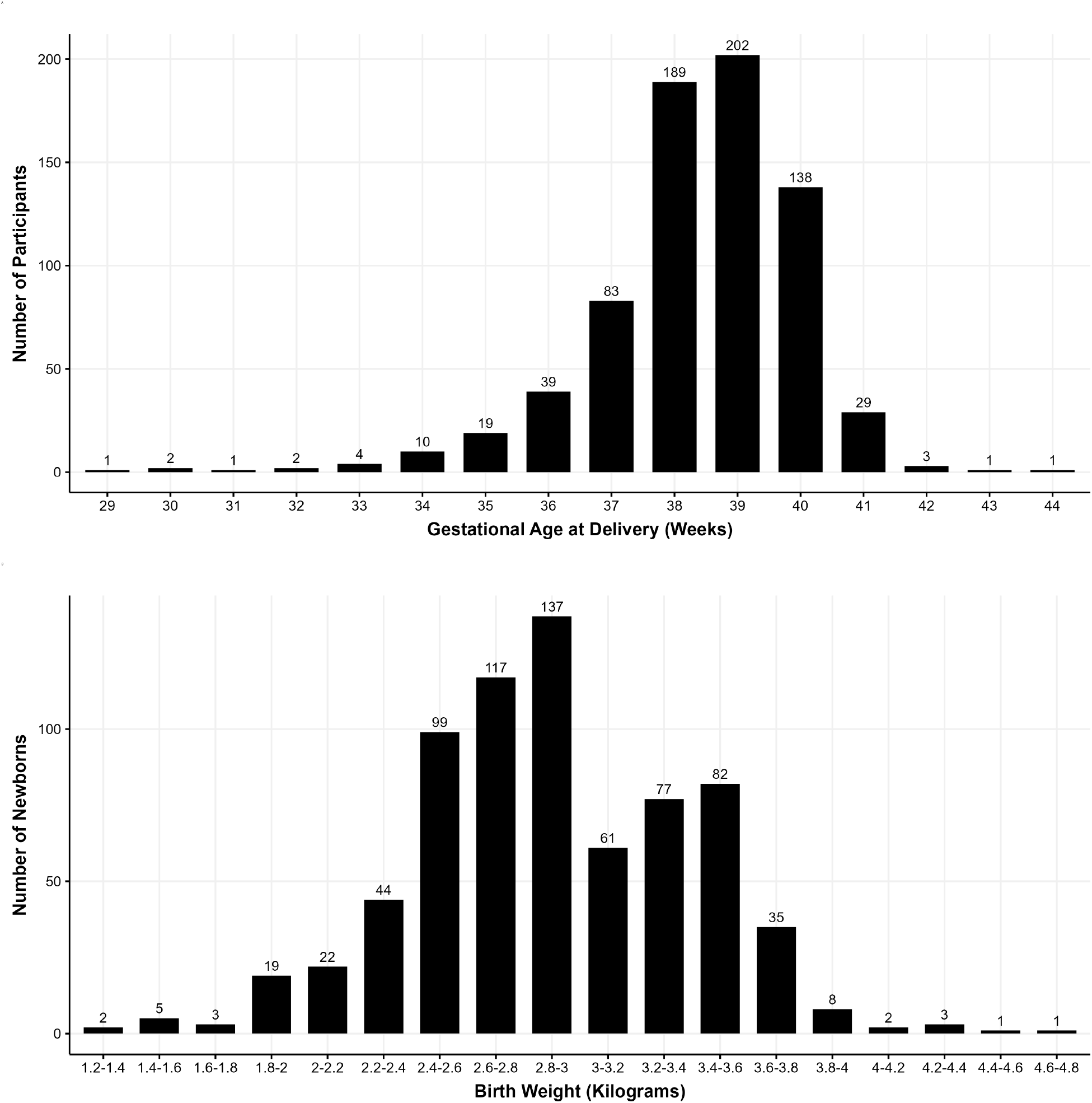
(A) Distribution of gestational age at delivery (weeks). (B) Distribution of birth weight at delivery (kilograms).

### Strengths and limitations

The prospective design of CORE is a strength, allowing standardised data collection and management harmonised with internationally recognised cohorts and thereby enabling pooled analyses and cross-cohort comparison. Rigorous quality-assurance measures were implemented to ensure the reliability and validity of the data, and the retention of both clean and caliper-annotated images supports validation of image-based models.

The study also has limitations. It was conducted at a single site drawing on a limited geographical catchment, which reduces the direct generalisability of the findings to other settings; like other single-site cohorts, its value for external validation is greatest when it is pooled with comparable cohorts rather than used in isolation. The population consisted primarily of hospital attendees rather than a community-based sample, which may limit representation of broader population characteristics. Models validated in CORE should therefore be expected to generalise most directly to urban, hospital-attending North Indian populations with similar socioeconomic and demographic characteristics; transportability to other LMIC settings should be assessed in dedicated external-validation studies. Another key limitation of our cohort is that, although the available sample size is sufficient to externally validate the discrimination of existing prediction models around an AUROC of 0.75 for SVN, SGA, and PTB, it is substantially smaller than recommended for precise assessment of calibration parameters Finally, the absence of continuous follow-up between 20 weeks and delivery limits the assessment of temporal change and longer-term outcomes.

### Collaboration

We welcome collaborators across academia, public health, and industry who wish to use CORE for external validation of risk-prediction models, secondary epidemiological analyses, or methodological work on AI in obstetric imaging. The consent obtained at recruitment restricts open public deposition of individual-level data and ultrasound images; access is therefore granted under a controlled-access model. Requests are submitted using a structured application form to the CORE Data Access Committee at THSTI, comprising the principal investigators, an external scientific reviewer, and an ethics representative. Each request is reviewed for scientific merit, ethical alignment with the original consent, and conflict-of-interest considerations; the expected turnaround from a complete application is six to eight weeks. Approved requests are executed under a Data Use Agreement covering permitted analyses, prohibition on re-identification, storage and destruction terms, and publication conditions. De-identified ultrasound images may be shared under the same agreement where the analysis falls within the scope of consent; image-level access for purposes outside that scope requires additional ethics-committee approval. Statistical code used for the analyses reported here is available from the corresponding author on reasonable request.

## Supporting information

STROBE_checklist

## Data Availability

De-identified clinical and ultrasound imaging data generated by the CORE cohort are available under a controlled-access model on reasonable request, subject to review and approval by the CORE Data Access Committee at THSTI and the execution of a Data Use Agreement, as described in the Collaboration section. Statistical code used for the analyses reported here is available from the corresponding author on reasonable request

## Funding

This work was supported by the CPH Early Career Fellowship from the DBT/Wellcome Trust India Alliance, grant number IA/CPHE/18/1.

Supporting institutions: Translational Health Science and Technology Institute (Faridabad, India); Institute of Biomedical Engineering, University of Oxford (Oxford, UK); and Hamdard Institute of Medical Sciences and Research and Hakeem Abdul Hameed Centenary Hospital (New Delhi, India).

## Author contributions

**RJ** contributed to writing the original draft of the manuscript, and **NS** performed the formal analysis. **AK** was responsible for methodology and contributed to writing, review, and editing. **NW** provided supervision, contributed to methodology, and participated in writing, review, and editing. **ReT** undertook data curation and contributed to writing, review, and editing. **AJ** also undertook data curation and contributed to writing, review, and editing. **SB** provided supervision, contributed to methodology, and participated in writing, review, and editing. **RcT** provided additional supervision, contributed to methodology, and participated in writing, review, and editing. **BKD** contributed to conceptualization and methodology, writing, review, and editing, and provided overall supervision as well as securing funding for the study.

## References

1 Lawn JE, Ohuma EO, Bradley E, et al. Small babies, big risks: Global estimates of prevalence and mortality for vulnerable newborns to accelerate change and improve counting. The Lancet. 2023;401:1707–19. doi: 10.1016/S0140-6736(23)00522-6

2 Saigal S, Doyle LW. An overview of mortality and sequelae of preterm birth from infancy to adulthood. The Lancet. 2008;371:261–9. doi: 10.1016/S0140-6736(08)60136-1

3 World Health Organization. SDG Target 3.2: End preventable deaths of newborns and children under 5 years of age. 2023.URL: https://www.who.int/data/gho/data/themes/topics/indicator-groups/indicator-group-details/GHO/sdg-target-3.2-newborn-and-child-mortality

4 Kim DW, Jang HY, Kim KW, et al. Design characteristics of studies reporting the performance of artificial intelligence algorithms for diagnostic analysis of medical images: Results from recently published papers. Korean Journal of Radiology. 2019;20:405–10. doi: 10.3348/kjr.2019.0025

5 Arun S, Grosheva M, Kosenko M, et al. Systematic scoping review of external validation studies of AI pathology models for lung cancer diagnosis. npj Precision Oncology. 2025;9:166. doi: 10.1038/s41698-025-00940-7

6 Johnson AEW, Pollard TJ, Shen L, et al. MIMIC-III, a freely accessible critical care database. Scientific Data. 2016;3:160035. doi: 10.1038/sdata.2016.35

7 Pollard TJ, Johnson AEW, Raffa JD, et al. The eICU collaborative research database, a freely available multi-center database for critical care research. Scientific Data. 2018;5:180178. doi: 10.1038/sdata.2018.178

8 Rockenschaub P, Akay EM, Carlisle BG, et al. External validation of AI-based scoring systems in the ICU: A systematic review and meta-analysis. BMC Medical Informatics and Decision Making. 2025;25:5. doi: 10.1186/s12911-024-02830-7

9 Siontis GCM, Tzoulaki I, Castaldi PJ, et al. External validation of new risk prediction models is infrequent and reveals worse prognostic discrimination. Journal of Clinical Epidemiology. 2015;68:25–34. doi: 10.1016/j.jclinepi.2014.09.007

10 Meertens LJE, Montfort P van, Scheepers HCJ, et al. Prediction models for the risk of spontaneous preterm birth based on maternal characteristics: A systematic review and independent external validation. Acta Obstetricia et Gynecologica Scandinavica. 2018;97:907– 20. doi: 10.1111/aogs.13358

11 Hadlock FP, Shah YP, Kanon DJ, Lindsey JV. Fetal crown-rump length: reevaluation of relation to menstrual age (5-18 weeks) with high-resolution real-time US. Radiology. 1992;182(2):501–505. doi:10.1148/radiology.182.2.1732970

12 Bhatnagar S, Majumder PP, Salunke DM, et al. A pregnancy cohort to study multidimensional correlates of preterm birth in india: Study design, implementation, and baseline characteristics of the participants. American Journal of Epidemiology. 2019;188:621–31. doi: 10.1093/aje/kwy284

13 Aftab F, Ahmed S, Ali SM, et al. Cohort profile: The alliance for maternal and newborn health improvement (AMANHI) biobanking study. International Journal of Epidemiology. 2021;50:1780–1781i. doi: 10.1093/ije/dyab124

14 Quinn JA, Munoz FM, Gonik B, et al. Preterm birth: Case definition & guidelines for data collection, analysis, and presentation of immunisation safety data. Vaccine. 2016;34:6047–56. doi: 10.1016/j.vaccine.2016.03.045

15 Villar J, Cheikh Ismail L, Victora CG, et al. International standards for newborn weight, length, and head circumference by gestational age and sex: The newborn cross-sectional study of the INTERGROWTH-21st project. The Lancet. 2014;384:857–68. doi: 10.1016/S0140-6736(14)60932-6

16 Vergouwe Y, Steyerberg EW, Eijkemans MJC, et al. Substantial effective sample sizes were required for external validation studies of predictive logistic regression models. Journal of Clinical Epidemiology. 2005;58:475–83. doi: 10.1016/j.jclinepi.2004.06.017

17 Riley RD, Debray TPA, Collins GS, et al. Minimum sample size for external validation of a clinical prediction model with a binary outcome. Statistics in Medicine. 2021;40:4230–51. doi: 10.1002/sim.9025

