## Supplementary material for "Cohort profile: the Cohort for Risk Prediction Model Evaluation (CORE) for external validation of models identifying high-risk pregnant women in the early second trimester": STROBE_checklist

STROBE Statement—checklist of items that should be included in reports of observational studies

|  | Item No. | Recommendation | Page  No. | Relevant text from manuscript |
| --- | --- | --- | --- | --- |
| **Title and abstract** | 1 | (*a*) Indicate the study’s design with a commonly used term in the title or the abstract | 1 | Title: Cohort profile ; Abstract: prospective cohort study. |
|  |  | (*b*) Provide in the abstract an informative and balanced summary of what was done and what was found | 1 | Abstract includes Purpose, Participants, Findings to date, and Future plans. |
| Introduction | | | |  |
| Background/rationale | 2 | Explain the scientific background and rationale for the investigation being reported | 2 | Introduction explains the scientific background and rationale, including the burden of neonatal mortality and the need for external validation of prediction models. |
| Objectives | 3 | State specific objectives, including any prespecified hypotheses | 2 | CORE was established to externally validate prediction models that identify high-risk pregnancies in the early second trimester. |
| Methods | | | |  |
| Study design | 4 | Present key elements of study design early in the paper | 3 | This is a prospective observational cohort study of pregnant women (N = 964) conducted at HIMSR, New Delhi, India, from August 2021 to March 2023. |
| Setting | 5 | Describe the setting, locations, and relevant dates, including periods of recruitment, exposure, follow-up, and data collection | 3 | Women were enrolled before 20 weeks of gestation and followed up at the 18–20 week scan and at delivery. |
| Participants | 6 | (*a*) *Cohort study*—Give the eligibility criteria, and the sources and methods of selection of participants. Describe methods of follow-up  *Case-control study*—Give the eligibility criteria, and the sources and methods of case ascertainment and control selection. Give the rationale for the choice of cases and controls  *Cross-sectional study*—Give the eligibility criteria, and the sources and methods of selection of participants | 3 | Inclusion criteria were age >18 years, a single live intrauterine pregnancy, gestational age estimated from crown–rump length measured before 14 weeks of gestation, and <20 weeks of gestation at enrolment. A total of 964 pregnant women who attended HIMSR for antenatal care, met the inclusion criteria, and provided consent were enrolled. |
|  |  | (*b*) *Cohort study*—For matched studies, give matching criteria and number of exposed and unexposed  *Case-control study*—For matched studies, give matching criteria and the number of controls per case | NA | Not applicable |
| Variables | 7 | Clearly define all outcomes, exposures, predictors, potential confounders, and effect modifiers. Give diagnostic criteria, if applicable | 5 | Outcomes and variables are defined for preterm birth, SGA, LGA, SVN, and the sociodemographic, clinical, obstetric, ultrasound, and delivery variables listed in Table 1. |
| Data sources/ measurement | 8* | For each variable of interest, give sources of data and details of methods of assessment (measurement). Describe comparability of assessment methods if there is more than one group | 4–5 | Clinical data were obtained from hospital records; structured sociodemographic and obstetric data were collected at enrolment; ultrasound at 18–20 weeks was performed by a trained radiologist; and delivery outcomes were documented. |
| Bias | 9 | Describe any efforts to address potential sources of bias | 4 | Potential bias was addressed through REDCap edit checks, manual monitoring, query resolution using source documents, and a two-tier ultrasound quality-assurance process with expert review of 10% of images monthly. |
| Study size | 10 | Explain how the study size was arrived at | 5 | The initial target was 800 participants based on an expected preterm-birth prevalence of approximately 13% and the rule of thumb of at least 100 outcome events; 164 additional participants were enrolled because loss to follow-up was higher than anticipated, giving a total of 964. |

Continued on next page

| Quantitative variables | 11 | Explain how quantitative variables were handled in the analyses. If applicable, describe which groupings were chosen and why |  |  |
| --- | --- | --- | --- | --- |
| Statistical methods | 12 | (*a*) Describe all statistical methods, including those used to control for confounding | 5 | The manuscript describes summary measures and reports prevalence estimates with 95% confidence intervals for principal outcomes. |
|  |  | (*b*) Describe any methods used to examine subgroups and interactions | NA | NA |
|  |  | (*c*) Explain how missing data were addressed | 5 | Missing data were not imputed; analyses used complete cases, and the number of participants with available data is given in the footnotes to Tables 2 and 3. |
|  |  | (*d*) *Cohort study*—If applicable, explain how loss to follow-up was addressed  *Case-control study*—If applicable, explain how matching of cases and controls was addressed  *Cross-sectional study*—If applicable, describe analytical methods taking account of sampling strategy | 6 | Outcomes were documented for 750 of 964 enrolled women. No further analytical method for loss to follow-up is described. |
|  |  | (*e*) Describe any sensitivity analyses | NA | No sensitivity analyses were reported. |
| Results | | | | |
| Participants | 13* | (a) Report numbers of individuals at each stage of study—eg numbers potentially eligible, examined for eligibility, confirmed eligible, included in the study, completing follow-up, and analysed | 6 | Of 964 enrolled women, 754 underwent an ultrasound scan at 18–20 weeks, and outcomes were documented for 750, comprising 23 abortions, 3 stillbirths, and 724 singleton live births. |
|  |  | (b) Give reasons for non-participation at each stage | NA | Reasons for non-participation at each stage were not reported in detail. |
|  |  | (c) Consider use of a flow diagram | 14 | Flow diagram is presented. |
| Descriptive data | 14* | (a) Give characteristics of study participants (eg demographic, clinical, social) and information on exposures and potential confounders | 6–7 | Participant characteristics are presented in Table 2, including demographic, clinical, social, and obstetric variables, along with information on exposures and potential confounders. |
|  |  | (b) Indicate number of participants with missing data for each variable of interest | 5, 7 | The number of participants with available data is given in the table footnotes, and complete-case analysis was used. |
|  |  | (c) *Cohort study*—Summarise follow-up time (eg, average and total amount) | NA | Not applicable |
| Outcome data | 15* | *Cohort study*—Report numbers of outcome events or summary measures over time | 6 | Outcome events are reported for preterm birth, abortion, stillbirth, low birth weight, SVN, and related newborn subgroups. |
|  |  | *Case-control study—*Report numbers in each exposure category, or summary measures of exposure | - | - |
|  |  | *Cross-sectional study—*Report numbers of outcome events or summary measures | - | - |
| Main results | 16 | (*a*) Give unadjusted estimates and, if applicable, confounder-adjusted estimates and their precision (eg, 95% confidence interval). Make clear which confounders were adjusted for and why they were included | NA | NA |
|  |  | (*b*) Report category boundaries when continuous variables were categorized | 5, 7 | Category boundaries are reported for BMI and for outcome categories such as preterm birth, SGA, and LGA using stated definitions. |
|  |  | (*c*) If relevant, consider translating estimates of relative risk into absolute risk for a meaningful time period | NA | NA |

Continued on next page

| Other analyses | 17 | Report other analyses done—eg analyses of subgroups and interactions, and sensitivity analyses |  |  |
| --- | --- | --- | --- | --- |
| Discussion | | | | |
| Key results | 18 | Summarise key results with reference to study objectives | 7 | The discussion summarises that CORE is a rigorously characterised cohort developed for external validation of prediction models in pregnancy. |
| Limitations | 19 | Discuss limitations of the study, taking into account sources of potential bias or imprecision. Discuss both direction and magnitude of any potential bias | 7 | Limitations discussed include single-site recruitment, limited geographical catchment, hospital-based sampling, limited precision for calibration assessment, and lack of continuous follow-up between 20 weeks and delivery. |
| Interpretation | 20 | Give a cautious overall interpretation of results considering objectives, limitations, multiplicity of analyses, results from similar studies, and other relevant evidence | 7 | The manuscript gives a cautious interpretation, presenting CORE as a useful LMIC-based resource whose value is greatest when pooled with comparable cohorts. |
| Generalisability | 21 | Discuss the generalisability (external validity) of the study results | 7 | The paper discusses generalisability mainly to urban, hospital-attending North Indian populations with similar socioeconomic and demographic characteristics, with transportability to other settings requiring separate assessment. |
| Other information | |  | | |
| Funding | 22 | Give the source of funding and the role of the funders for the present study and, if applicable, for the original study on which the present article is based | 8 | This work was supported by the CPH Early Career Fellowship from the DBT/Wellcome Trust India Alliance, grant number IA/CPHE/18/1. |

*Give information separately for cases and controls in case-control studies and, if applicable, for exposed and unexposed groups in cohort and cross-sectional studies.

**Note:** An Explanation and Elaboration article discusses each checklist item and gives methodological background and published examples of transparent reporting. The STROBE checklist is best used in conjunction with this article (freely available on the Web sites of PLoS Medicine at http://www.plosmedicine.org/, Annals of Internal Medicine at http://www.annals.org/, and Epidemiology at http://www.epidem.com/). Information on the STROBE Initiative is available at www.strobe-statement.org.
